# Prediction of Buruli ulcer treatment shortening with novel beta-lactam-containing antimicrobial combinations

**DOI:** 10.64898/2026.02.28.26347324

**Authors:** Umberto Villani, Salvatore D’Agate, Emma Sáez López, Santiago Ramón-García, Oscar Della Pasqua

## Abstract

**Introduction:** Buruli ulcer (BU) is a neglected tropical disease primarily affecting skin and sometimes bone. Standard therapy consists of rifampicin (RIF, once daily) plus clarithromycin (CLA, twice daily) over 8 weeks. Adding amoxicillin-clavulanate (AMX/CLV) may shorten treatment, but predicting treatment success before clinical trial implementation is challenging.

**Aims:** To assess the probability of bacterial eradication following treatment with novel investigational BU regimens over different intervals using a mechanism-based modelling and simulation approach.

**Methods:** *In vitro* time-kill assays with RIF, CLA, and AMX/CLV alone and in combination were performed with a range of clinical isolates of *Mycobacterium ulcerans*. Bactericidal activity was characterized using a bacterial growth dynamics model, including an Emax function to describe the drug effect. Subsequently, clinical trial simulations were performed to evaluate drug disposition and skin penetration in a cohort of virtual subjects, taking into account interindividual variability in pharmacokinetics and pharmacodynamics (n=70/arm). Several regimens, including standard therapy and AMX/CLV-containing combinations with higher RIF doses were assessed. The probability of eradication at 4–8 weeks was assessed across strains with different susceptibility and assuming varying bacterial load at start of treatment.

**Results:** Beta-lactam containing combinations resulted in higher potency and maximum killing rates relative to the currently recommended regimens. Consequently, regimens containing AMX/CLV with higher RIF doses (20 mg/kg q.d. or 10 mg/kg b.i.d.) outperformed standard therapy, achieving 100% eradication within 4 weeks for baseline loads up to 1,000 CFU/mL across most isolates, except one from China. At higher loads (10,000 CFU/mL), 6 weeks were required.

**Conclusions:** The use of mechanism-based modelling and clinical trial simulations provides a robust translational framework for the evaluation of novel therapies for neglected diseases, such as BU. Irrespective of differences in bacterial susceptibility, adding AMX/CLV or using RIF–AMX/CLV dual therapy may reduce BU treatment from 8 to 4 weeks.

## 1. Introduction

Buruli ulcer (BU) is a neglected tropical disease (NTD) caused by *Mycobacterium ulcerans* that mainly affects the skin and sometimes the bones, leading to ulcers and permanent disfigurement in absence of appropriate treatment^1^. More than 80% of the global burden falls in Africa, with nearly 50% being children under the age of 15 years^2^, although it is also endemic in other parts of the world, such as Australia. Historically, BU management relied exclusively on surgical excision of lesions followed by skin grafting. While this approach was effective, it was associated with prolonged hospitalization, high treatment costs, and recurrence rates of approximately 15%^3,4^. A significant milestone in BU treatment occurred in 2004 with the introduction of an antimicrobial regimen combining oral rifampicin (RIF) and intramuscular streptomycin (STR), which improved cure rates and reduced the need for amputation, but caused unacceptable levels of ototoxicity and nephrotoxicity^5,6^. Since 2017, an all-oral antimicrobial regimen consisting of RIF (10 mg/kg q.d.) and CLA (7.5 mg/kg b.i.d.) for 8 weeks is recommended by the World Health Organization (WHO)^7^. In addition, complementary treatments are needed for an optimal recovery, including wound care and surgery, as well as physiotherapy in severe cases. Despite this progress, adherence to the current treatment still poses challenges since this disease primarily impacts people in rural and resource-limited settings, where access to health services is challenging. Therefore, a shortened and highly effective all-oral regimen could improve the care of BU patients.

One promising avenue for BU treatment shortening involves the addition of amoxicillin/clavulanate (AMX/CLV) to the current standard of care (RIF+CLA). The rationale for this inclusion was supported by *in vitro* investigations demonstrating strong synergistic activity between RIF and AMX^8,9^, along with its excellent safety profile both in adults and children and its widespread availability worldwide. The potential of this triple-antimicrobial combination to shorten the treatment to 28 days is currently being assessed in two ongoing clinical trials that evaluate its non-inferiority to the standard of care (https://blms4bu.org/) (NCT05169554, PACTR202209521256638)^10^. Notably, the strong synergy between RIF and AMX/CLV has been predicted to enhance the probability of target attainment (PTA) of BU treatments incorporating this combination in the clinic^11^, allowing for twice-daily administration of AMX, thereby potentially enhancing adherence. However, that investigation did not assess whether these regimens warrant bacterial eradication in BU patients within four weeks, which would ultimately determine the clinical impact of adding AMX to the standard of care.

Mechanism-based drug-disease modelling and simulation (M&S) has been shown to be an effective tool to optimise the development of investigational treatment regimens in many therapeutic areas^12^, including tuberculosis (TB)^13,14^. As such, to date it has gained wide recognition from drug developers and regulatory agencies^15,16^. More specifically, the use of clinical trial simulations (CTS) allow for *in silico* implementation of a trial, providing insight into the probability of pharmacological success before implementing a clinical study with real patients. Despite the merit of the approach for establishing the dose rationale in humans and optimising study protocol design, the use of these techniques is largely unexplored for the evaluation of novel antimicrobial therapies for BU.

Within this context, the aims of this study were: (i) to develop a translational framework that integrates *in vitro* experimental data with *in silico* drug-disease modelling and simulation to predict the probability of bacterial eradication in patients affected by BU, taking into account interindividual variability in pharmacokinetics and differences in the susceptibility of clinical isolates to selected drugs, (ii) to assess whether adding AMX/CLV to the current standard of care warrants bacterial eradication within a reduced treatment course of 28 days, (iii) to investigate the potential of alternative regimens with RIF and AMX/CLV to further shorten or simplify BU treatment, which have not been considered for the evaluation of efficacy in ongoing beta-lactam containing BU trials, including substitution of CLA with AMX/CLV and the use of higher RIF doses, as supported by TB studies demonstrating enhanced mycobacterial killing and adequate safety profiles at elevated dosing levels^17^.

## 2. Methods

### 2.1. Experimental data

*In vitro* time-kill assay (TKA) data of five different *M. ulcerans* clinical isolates with different geographical provenance, each one treated with different drug/drug combinations was available for this analysis^9^. The following isolates were included: ITM 000932, ITM 941327 and ITM C05142 from Australia, ITM 070290 from China, and ITM C08756 from Japan. Bacterial load was quantified at multiple time points using the BacTiter-Glo™ Microbial Cell Viability Assay (Promega, WI, USA), which produces an ATP-based luminescent signal proportional to the number of viable bacterial cells (relative light units, RLU). Briefly, cultures were first incubated for three days to reach the exponential growth phase prior to the start of treatment. On Day 0 drugs were added at different concentrations, and bacterial load was subsequently assessed longitudinally over a period of 28 days (i.e., on days 0, 1, 3, 7, 10, 14, 21, and 28 post-treatment initiation). RIF, CLA, and AMX were tested individually as monotherapies at seven concentration levels, chosen as fold-changes from reference minimum inhibitory concentration (MIC) values, which were 0.1 μg/mL, 0.25 μg/mL and 0.5 μg/mL, respectively. The tested concentrations covered the following fold-MIC values: 0.05×, 0.25×, 1×, 4×, 10×, 20×, and 100×. In the case of combination experiments, two matching nominal concentration levels were used—0.25× MIC and 1× MIC. Further assay details are described within Sáez-López *et al*^9^.

### 2.2. Drug-disease modelling and overall translational framework

An outline of the steps required to implement a translational framework to predict the probability of bacterial eradication and corresponding treatment performance in clinical settings is depicted in **Figure *1***. Briefly, a bacterial growth dynamics model was used to describe the *M. ulcerans* growth profile in time-kill assays over 28 days in the absence of drugs. Pharmacokinetic-pharmacodynamic (PKPD) relationships were then parameterised to describe the antibacterial activity of RIF in monotherapy and in combinations with CLA and AMX/CLV. This drug-disease model was used to predict the time course of antibacterial activity in a virtual cohort of patients, under the assumption of comparable growth dynamics and PKPD relationships *in vivo*. Systemic exposure to the selected drugs and drug combinations was derived from previously published pharmacokinetic models, taking into account known clinical and demographic characteristics of patients with BU. Subsequently, clinical trial simulations featuring several arms were implemented to establish whether any given treatment resulted in bacterial eradication within 28 days from its initiation. Model parameterisation, estimation and simulation steps were implemented using a nonlinear-mixed effects modelling approach.

**Figure 1:**
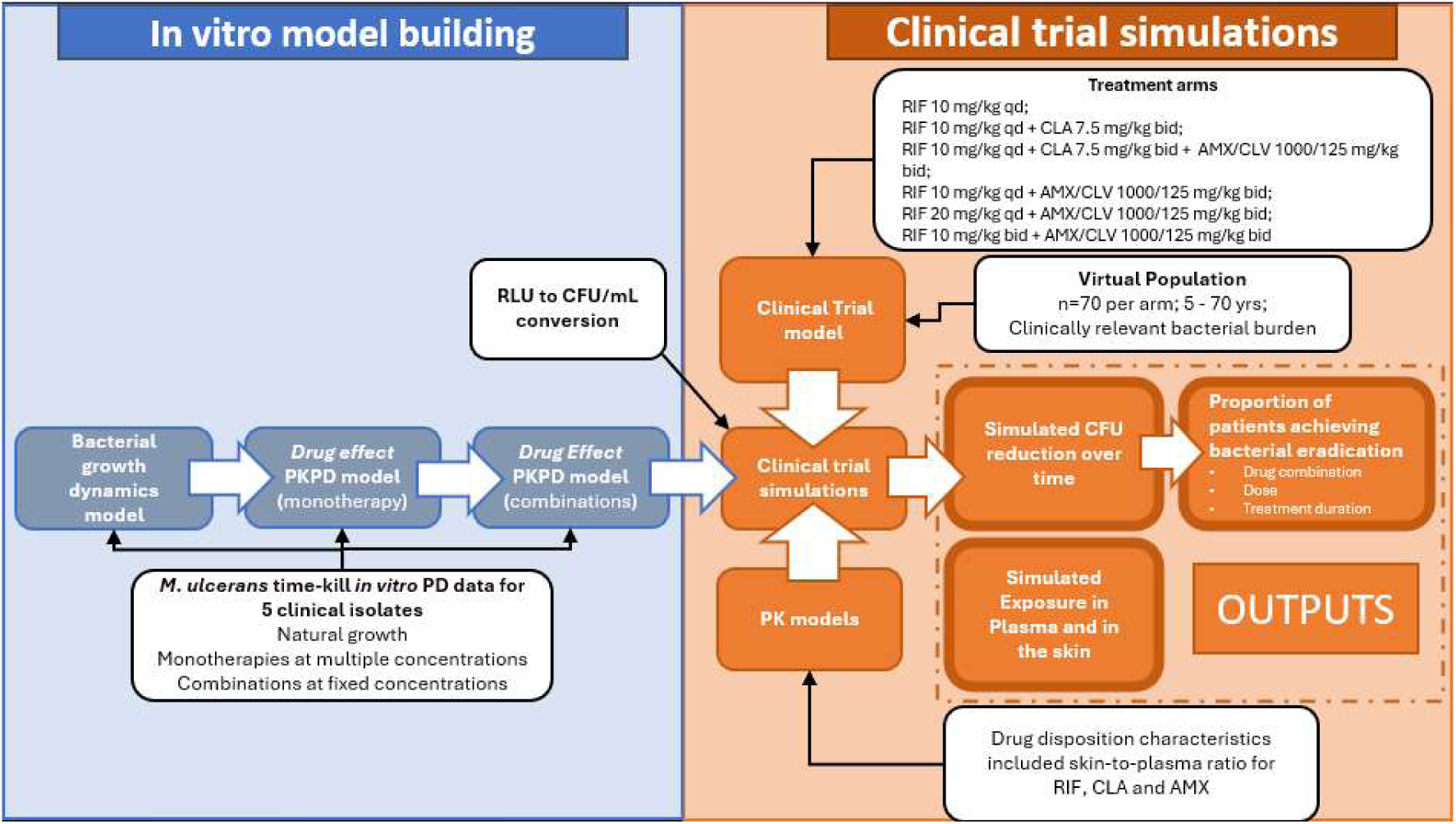
Flowchart describing the translational framework for the prediction of bacterial eradication in patients affected by BU. Abbreviations: AMX/CLV = amoxicillin/clavulanate, b.i.d. = twice daily, CLA = clarithromycin, PKPD = pharmacokinetic-pharmacodynamic, q.d. = once daily, RIF = rifampicin, RLU = relative light units, CFU = colony forming units.

### 2.3. Model-based characterisation of the *in vitro* efficacy of antimicrobial regimens in *M. ulcerans* time-kill assays

Initially, *in vitro* data in the absence of treatment was used to characterise the natural growth of different clinical isolates of *M. ulcerans*. Different models were evaluated for their ability to fit the experimental data at hand. Initially, only the Verhulst (logistic) and Gompertz models were considered^18^. However, exploratory data analyses revealed oscillations in bacterial load near the carrying capacity of the *in vitro* system, supporting the inclusion of the Hutchinson model^19^ (i.e., a delay logistic model). Structural model parameters were estimated separately for each bacterial isolate.

In a subsequent step, experimental data including treatment with RIF monotherapy were used to characterize the concentration–effect relationship of RIF. The approach assumes the use of RIF as the backbone drug in combination regimens. The parameterisation of concentration-dependent bacteriostatic/bactericidal effects of RIF on *M. ulcerans* was attempted both as a multiplicative reduction to the bacterial growth rate or as first-order killing rate. Several pharmacodynamic (PD) structural models were tested, including log-linear, Emax and sigmoidal Emax models^20^. Based on initial model-building iterations, a time lag factor on the onset of the antibacterial effect was required to describe the observed time course of the effect. PKPD parameter estimates were derived individually for each bacterial strain. Drug degradation in the culture medium over the course of the experiments was assumed to follow exponential decay from nominal concentrations, with an approximate half-life of 6 days^21^.

Building on the parameterization of bacterial growth dynamics of *M. ulcerans* and monotherapy effects of RIF, the addition of companion drugs was modelled as a shift in the estimates of the PKPD parameters describing the antibacterial activity of the backbone drug, namely EC_50_ and E_max_, as previously proposed for the characterisation of antitubercular drug regimens^22^. This approach allows for a semi-mechanistic interpretation of estimates of drug-specific parameters, while retaining model robustness and minimising identifiability issues.

Specifically, under the assumption that RIF achieves maximal killing rates at sufficiently high concentrations, E_max_ was considered to remain unchanged and the effect of CLA and AMX/CLV was parameterised as a shift to RIF’s potency (i.e., EC_50_). Similarly, in case of the triple antimicrobial regimen, the shift for the addition of AMX/CLV was calculated taking as reference the apparent EC_50_ estimated from the RIF + CLA standard regimen.

The decrease in the objective function value (OFV) by at least 3.84 (α = 0.05, based on a χ^2^ test with 1 degree of freedom) between nested models was used to assess the statistical significance of added model complexity. Goodness-of-fit plots (population/individual predicted *vs.* observed concentrations, observed concentrations /time *vs*. conditional weighted residual) were assessed at each step of the model building process. In addition, visual predictive checks (VPCs) were used to assess the predictive performance of the final model

#### 2.3.1. Translational assumptions

Following the implementation of the growth dynamics model, and characterisation of the concentration-effect relationships of different drug combinations, model parameter estimates describing bacterial growth and PKPD properties were applied to clinical settings using simulation scenarios, taking into consideration several translational assumptions. First, as the PKPD model was developed using *in vitro* data where bacterial viability was assessed via a luminescence-based readout (relative light units, RLU), it was necessary to establish a relationship between the two bacterial viability biomarkers – CFU and RLU - in this particular experimental context. Notably, this correlation has been shown to be linear by multiple studies^9,23,24^. A linear regression model was subsequently implemented and integrated in the translational parametrisation to convert simulated RLU values into CFU values (**Supplementary material, Appendix A**).

Subsequently, for drug-specific parameters, available data on human plasma protein binding and the skin-to-plasma exposure ratio (**Table S*2***) were used to ensure accurate description of the pharmacokinetics at the site of infection. Additionally, given the established mechanism of action of RIF^25^, it was assumed that steady-state concentration (Css), rather than instantaneous concentrations drive its bactericidal effect. Likewise, Css of companion drugs was considered to drive the observed shift in RIF potency following the administration of different drug combinations, in line with the strategy illustrated in **Figure *2***. For disease-related parameters, as the median incubation time in patients has previously been estimated to be around 135 days^26^, it was assumed that at the time of diagnosis and start of the treatment, the infection was well-established and had reached equilibrium in terms of bacterial burden within the lesion. Therefore, to explore the implications of varying levels of infection severity on antimicrobial treatment duration, different bacterial load values were considered at baseline (10-10^4^ Colony Forming Units, CFU/mL), which reflect a clinically plausible range^27^.

**Figure 2:**
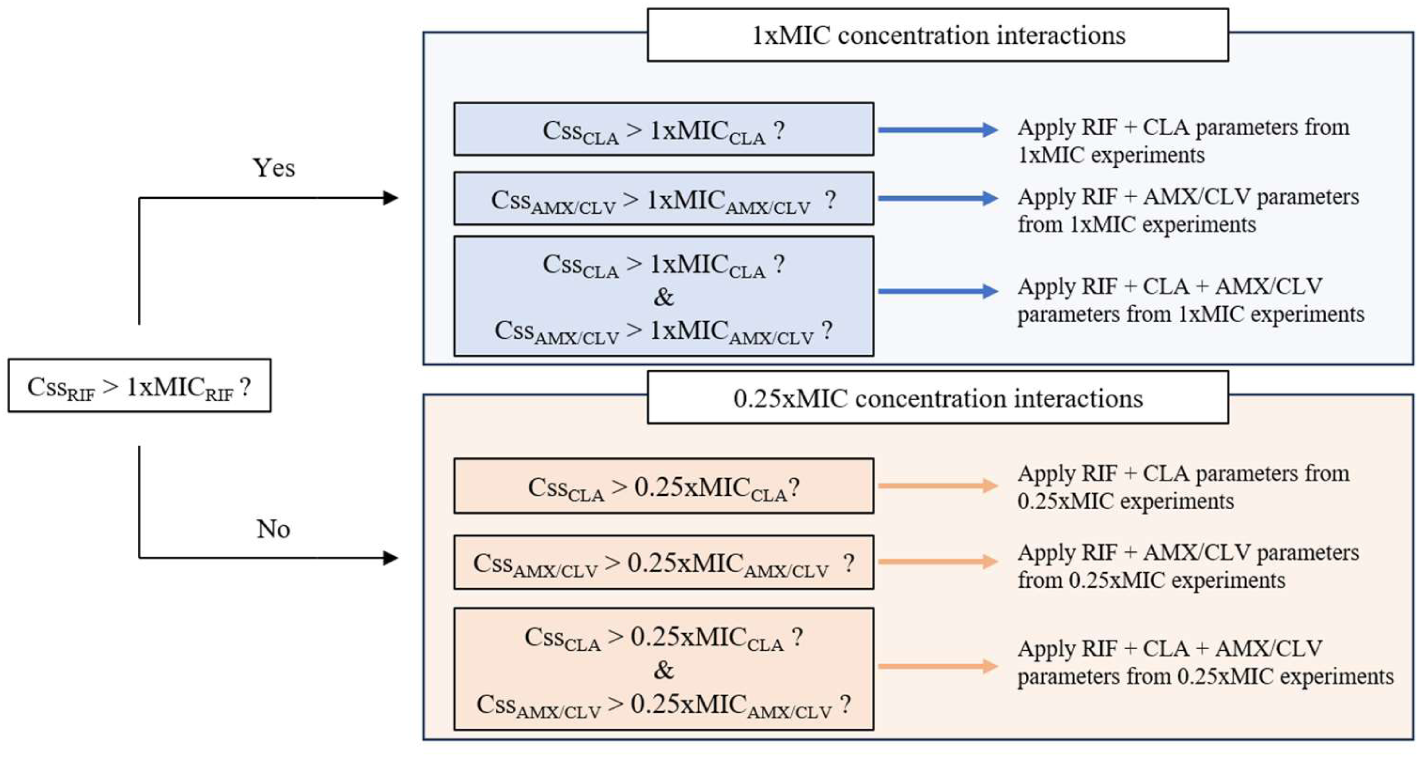
Overview of the strategy for incorporating pharmacodynamic interactions into clinical trial simulations. As experimental data were limited to discrete steps and ratios between drugs, assumptions regarding the magnitude of the effect of combination regimens were required to handle the continuous nature of pharmacokinetic data *in vivo* in a clinical setting. Parameters describing the drug-disease model for combinations are applied in simulations based on how steady state concentrations (C_ss_) of the three antimicrobials relate to the tested *in vitro* experimental conditions (i.e., 0.25xMIC and 1xMIC conditions, respectively). When the C_ss_ of rifampicin (RIF) exceeded the 1×MIC threshold, interaction parameter estimates derived from *in vitro* experiments at the 1×MIC level were used—provided that the C_ss_ of the companion drugs also exceeded this threshold. Similarly, when the C_ss_ of RIF was below the 1×MIC threshold, the interaction parameters derived from *in vitro* experiments at the 0.25×MIC level were used - provided that the C_ss_ of the companion drugs also exceeded this threshold. If these criteria were not met, then estimates of the antibacterial activity of RIF monotherapy were applied, i.e., it was assumed that pharmacodynamic interaction was not detectable.

### 2.4. Clinical trial simulations

#### 2.4.1. Virtual patient cohorts

Given the ongoing clinical trials in Benin, Côte d’Ivoire and Togo (the BLMs4BU trial^10^, *NCT05169554*, *PACTR202209521256638*), a virtual patient cohort was used that reflect our previous analysis evaluating the PTA of antimicrobial regimens in Buruli Ulcer^11^. The virtual cohort included 70 paediatric and adult patients per treatment arm and was designed to reflect realistic baseline and demographic characteristics - such as age, body weight, body mass index (BMI), and creatinine clearance. BU incidence data from the WHO^2,28^ were used to replicate realistic age groups for recruitment, estimating that nearly half (48%) of cases occur in children under 15 years of age. The paediatric subgroup of the virtual population reflected growth parameters from a previous cross-sectional study of children, aged 6–18 years, in Calabar, Nigeria^29^ (**Table S*1***).

#### 2.4.2. Simulated treatment arms and endpoints

Simulation scenarios were implemented to evaluate the pharmacokinetics and pharmacodynamic profiles of the following treatment regimens (**Table 1**): (i) RIF (q.d., 10 mg/kg); (ii) RIF (q.d., 10 mg/kg) + CLA (b.i.d., 7.5 mg/kg); (iii) RIF (q.d., 10 mg/kg) + CLA (b.i.d., 7.5 mg/kg) + AMX/CLV (b.i.d., 1000/125 mg/kg); (iv) RIF (q.d., 10 mg/kg) + AMX/CLV (b.i.d., 1000/125 mg/kg); RIF (q.d., 20 mg/kg) + AMX/CLV (b.i.d., 1000/125 mg/kg); (vi) RIF (b.i.d., 20 mg/kg) + AMX/CLV (b.i.d., 1000/125 mg/kg).

**Table 1:** Dosing regimen investigated in clinical trial simulations. Abbreviations: RIF = rifampicin, CLA = Clarithromycin, AMX/CLV = Amoxicillin/clavulanate, qd = once daily, bid = twice daily.

| Scenario name | Drugs, doses and dosing regimens | Note |
| --- | --- | --- |
| **RIF** | RIF (q.d., 10 mg/kg) | Hypothetical scenario evaluating RIF alone at standard dosage. |
| **RIF+CLA** | RIF (q.d., 10 mg/kg) + CLA (b.i.d., 7.5 mg/kg); | WHO-recommended dosing regimen, standard of care. |
| **RIF+CLA+AMX/CLV** | RIF (q.d., 10 mg/kg) + CLA (b.i.d., 7.5 mg/kg) + AMX/CLV (b.i.d., 1000/125 mg/kg) | Regimen proposed for NCT05169554 and PACTR202209521256638 studies |
| **RIF+AMX/CLV** | RIF (q.d., 10 mg/kg) + AMX/CLV (b.i.d. 1000/125 mg/kg) | Regimen proposed for NCT05169554 and PACTR202209521256638 studies, without CLA. |
| **RIF20qd+AMX/CLV** | RIF (q.d., 20 mg/kg) + AMX/CLV (b.i.d., 1000/125 mg/kg) | Regimen proposed for NCT05169554 and PACTR202209521256638 studies, without CLA. RIF dose is doubled |
| **RIF10bid+AMX/CLV** | RIF (b.i.d., 10 mg/kg) + AMX/CLV (b.i.d., 1000/125 mg/kg). | Regimen proposed for NCT05169554 and PACTR202209521256638 studies, without |
|  |  | CLA. RIF dose is doubled and given bid, to match AMX/CLV dosing schedule. |

Previously published pharmacokinetic models of RIF, CLA and AMX were used to simulate clinical concentration-time profiles of the investigated drugs^11^. CLV was assumed to remain pharmacologically active throughout the treatment course at the administered dose, regardless of potential intra- and inter-individual variability (IIV) in its exposure. Therefore, individual concentration–time profiles for CLV were not simulated.

Individual trajectories of the bacterial burden, expressed as CFU/mL, were simulated over an 8-week treatment period. Bacterial eradication for each individual was considered achieved when simulated CFU/mL values decreased below 1. The cumulative proportion of patients reaching bacterial eradication at each time point was subsequently calculated across the treatment period.

Simulations of individual Css values for all antimicrobials were also performed to assess the impact of interindividual variability in skin exposure.

### 2.5. Software

All modelling and simulation analyses were conducted in NONMEM v7.5 (Icon Development Solutions, Ellicott City, MD, USA) and Ps*N* (*Perl-speaks-*NONMEM, University of Uppsala, Sweden). During model building, parameter estimation was conducted using the first-order conditional estimation method with the interaction option (FOCE-I). Data manipulation, including the creation of graphical and statistical summaries was performed in R v.4.5.0 (R Foundation for Statistical Computing, Vienna, Austria).

## 3. Results

### 3.1. Drug-disease modelling of the antibacterial activity of drug combinations against M. ulcerans

The growth dynamics of *M. ulcerans* and antibacterial effect of RIF and selected drug combinations observed in *in vitro* in time-kill assays was parameterised in a way that enables us to disentangle drug-specific from disease-related properties, and consequently provides the basis for subsequent use of the model for translational purposes. For the sake of clarity, this process involved: (i) parameterisation of the *in vitro* natural growth dynamics of *M. ulcerans*, (ii) evaluation of the monotherapy antibacterial activity of RIF (the backbone drug of combination regimens) and, (iii) determination of how the RIF concentration-effect relationship shifted by the addition of companion drugs, either in pairwise manner (RIF+CLA; RIF+AMX/CLV) or as triple combinations (RIF+CLA+AMX/CLV). The natural growth dynamics and the monotherapy concentration-effect curve of RIF were best described by a delayed logistic model and an Emax model parameterising the first-order bacterial killing rate, incorporating a time lag to account for the delayed onset of the antibacterial effect after start of treatment. **Table *2*** and **Figure S*2*** show respectively the parameter estimates and the goodness-of-fit (GoF) plots for the final monotherapy drug-disease model.

**Table 2:**
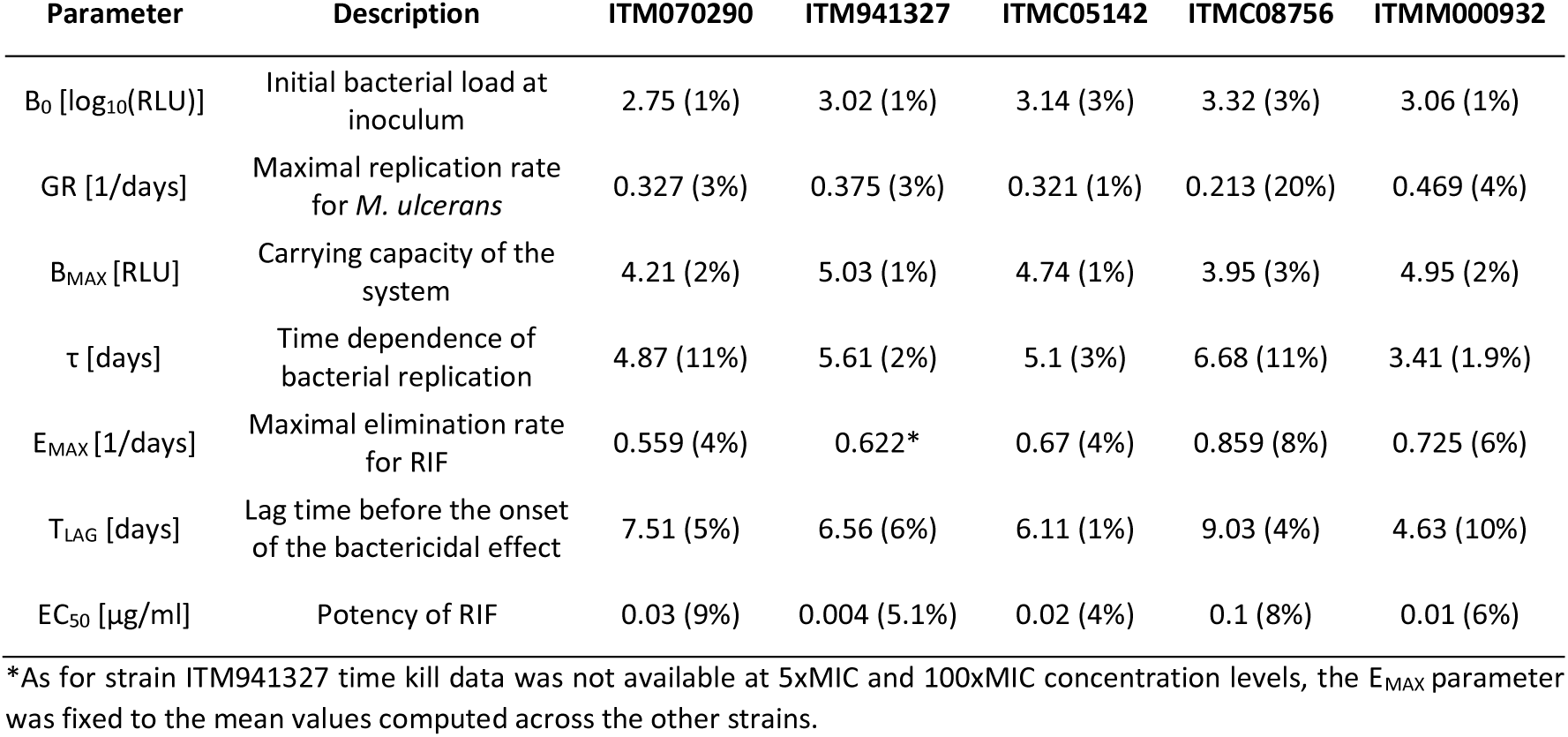
Parameter values and precision for the drug-disease model describing the bacterial growth dynamics of five different *Mycobacterium ulcerans* clinical isolates and the concentration-effect relationship of rifampicin. The precision of parameter estimates is shown as coefficient of variation (CV%). Abbreviations: RLU = relative light units, RIF = rifampicin.

Structural parameters were estimated with good precision and the model showed no evidence of systematic bias across all the graphical diagnostics. Notably, bacterial replication rates, as well as maximal elimination rates for RIF were similar across the different strains. However, different susceptibilities of RIF were noted, and mechanistically captured as variations in the potency of RIF, which varied almost 100-fold across different isolates. Finally, visual predictive checks (VPCs) performed on the final parameterisation showed that model predictions reproduced the observed experimental data across all evaluated conditions (**Figure *3***).

**Figure 3:**
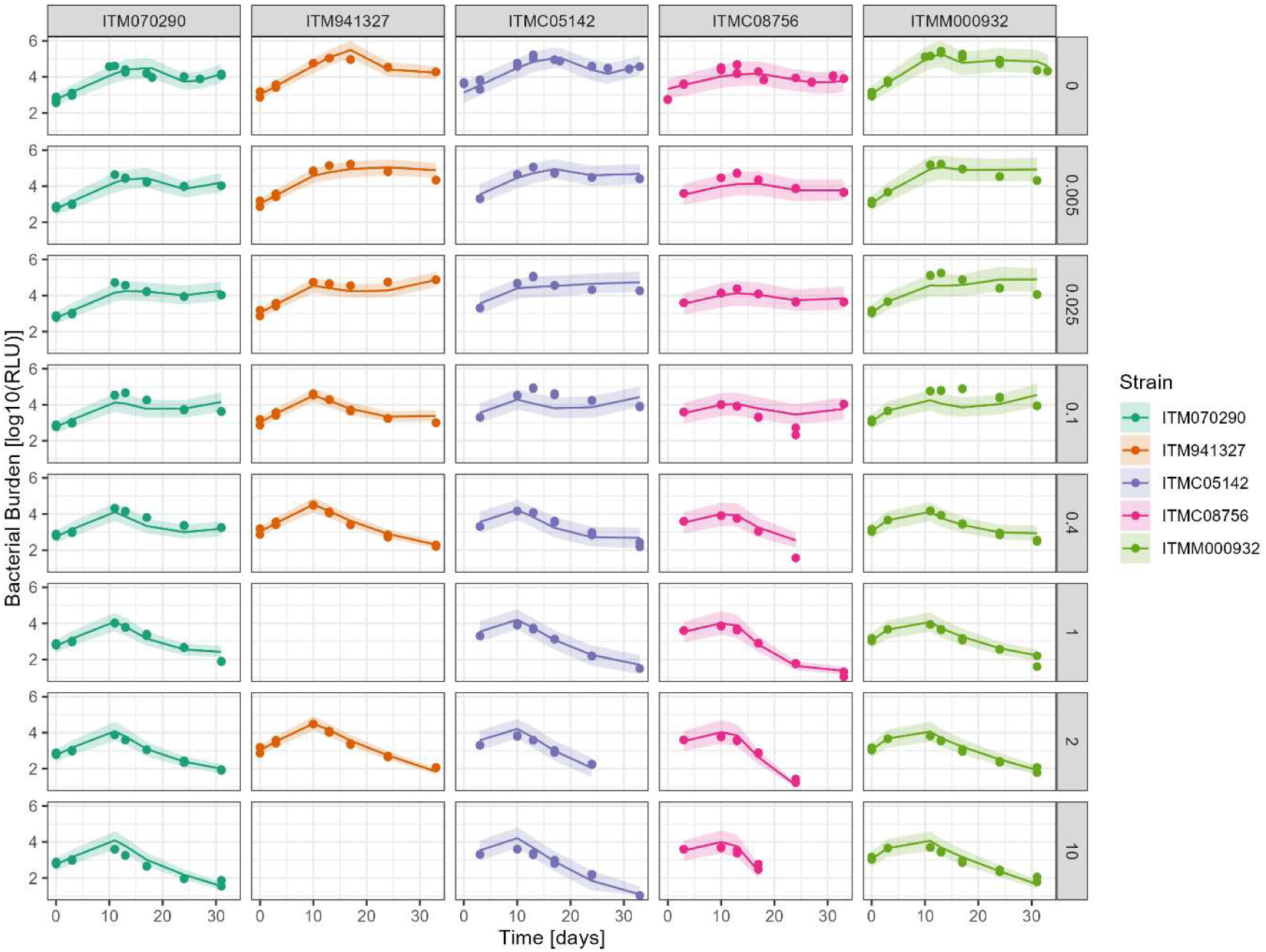
Visual predictive checks for the drug-disease model describing the bacterial growth dynamics of five different *Mycobacterium ulcerans* clinical isolates and the concentration-effect relationship of rifampicin. Columns represent model predictions and experimental data for each bacterial strain, while rows represent increasing rifampicin concentrations [μg/mL]. In each plot, dots represent the experimental data, solid lines represent the median model prediction and the shaded areas represent the 5^th^-95^th^ prediction interval of the model.

Time-kill data of antimicrobial combinations at varying concentration levels was found to be adequately described as a shift in the potency of RIF. Notably, accurately fitting the experimental data required introducing a multiplicative factor to account for the lag time in the onset of detectable antibacterial activity between RIF monotherapy and combination therapies. **Table *3*** and ***Figure S3*** show respectively the parameter estimates of the drug effect of combinations for each isolate and the goodness-of-fit plots of the final combination model. Consistent with monotherapy estimation procedures, model predictions demonstrated no systematic bias, and the combination effect parameters were estimated with acceptable precision. Overall, the addition of AMX/CLV to RIF, alone or with CLA, resulted in an increased bactericidal effect, denoted by higher *apparent* potencies (i.e., lower EC_50_ values) for AMX/CLV-containing regimens. This was especially evident in experimental conditions where nominal drug concentrations approached 0.25xMIC values (**Figure 4**). In fact, increasing nominal concentration of all drugs from 0.25xMIC to 1xMIC did not lead to an appreciable increase in the overall bactericidal effect, suggesting that AMX/CLV-containing antimicrobial combinations at 0.25xMIC concentrations already reach the expected maximum killing rate of RIF.

**Figure 4:**
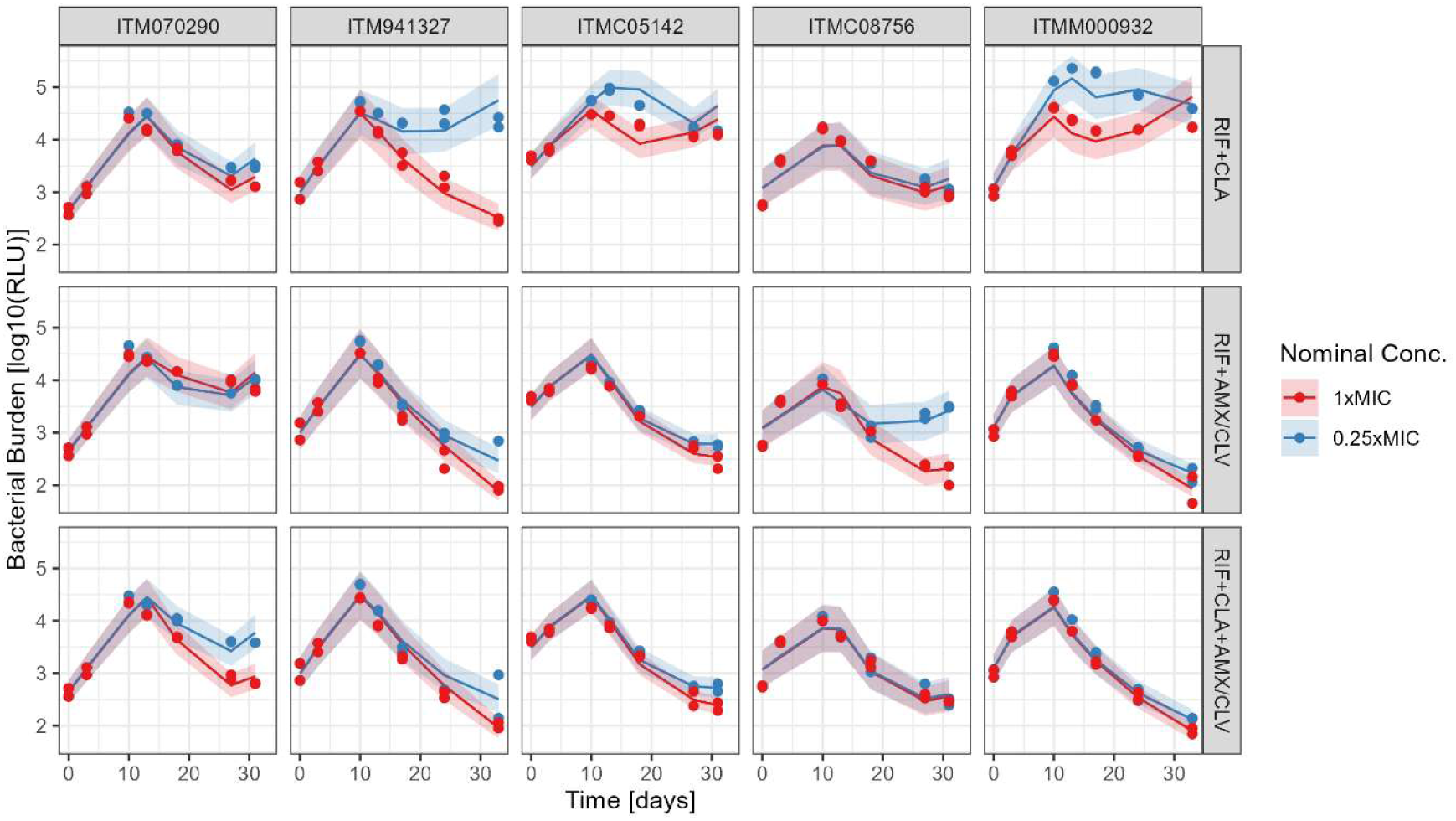
Visual predictive checks of the drug-disease model describing the bactericidal effect of investigated drug combinations (RIF+CLA, RIF+AMX/CLV and RIF+CLA+AMX/CLV) against five different clinical isolates. Columns represent model predictions and experimental data for each bacterial strain, while rows represent different drug combinations. Different colours depict longitudinal data and model predictions for the two available nominal concentration levels. In each plot, dots represent the experimental data, solid lines represent the median model prediction and shaded areas represent the 5^th^-95^th^ prediction interval of the model.

**Table 3:** Estimated shifts in the potency of RIF when companion drugs are added in *in vitro* experiments (CLA, AMX/CLV and CLA+AMX/CLV) at different concentration levels. Results are shown for all the clinical isolates included in this study. Precision of parameter estimates is shown as coefficient of variation (CV%). For clarity, the estimated monotherapy potency values for rifampicin are provided in the table. Abbreviations: RIF = rifampicin, CLA = clarithromycin, AMX/CLV = amoxicillin/clavulanate.

| Parameter | ITM070290 | ITM941327 | ITMC05142 | ITMC08756 | ITMM000932 |
| --- | --- | --- | --- | --- | --- |
| $EC_{50_{RIF}}$ [µg/ml] | 0.03 | 0.004 | 0.02 | 0.1 | 0.01 |
| **0.25xMIC shifts<sup>a</sup>** |  |  |  |  |  |
| $\Theta_{EC50_{CLA}}$ | 0.133 (7.70%) | 0.886 (1.50%) | 37.2 (56.20%) | 0.1 (0.60%) | 12.6 (1.4%) |
| $\Theta_{EC50_{AMX/CLV}}$ | 0.238 (5.30%) | 0.0843 (1.70%) | 0.0726 (0.20%) | 0.127 (0.20%) | 0.027 (0.6%) |
| $\Theta_{EC50_{CLA+AMX/CLV}}^b$ | 1.35 (1.40%) | 0.104 (0.20%) | 0.00186 (55.90%) | 0.556 (0.10%) | 0.002 (18.2%) |
| **1xMIC shifts<sup>a</sup>** |  |  |  |  |  |
| $\Theta_{EC50_{CLA}}$ | 0.327 (7.40%) | 0.368 (1.80%) | 1.32 (0.50%) | 0.348 (0.50%) | 1.19 (1.4%) |
| $\Theta_{EC50_{AMX/CLV}}$ | 1.65 (11.70%) | 0.0663 (5.70%) | 0.225 (0.20%) | 0.182 (0.60%) | 0.0668(0.8%) |
| $\Theta_{EC50_{CLA+AMX/CLV}}^b$ | 0.616 (0.60%) | 0.26 (2.40%) | 0.146 (0.50%) | 0.612 (0.10%) | 0.0519 (0.7%) |
| $\epsilon_{add}^2$<sup>c</sup> | 0.0025 (3.9%) | 0.00387 (17.2%) | 0.0017 (15.9%) | 0.0049 (6.5%) | 0.0026 (17.2%) |

<sup>a</sup>:Fluctuations in the time to onset of effect (TLAG) were observed in several combination experiments. This variation was estimated to range from 40% to 140% of the original value and was attributed to experimental variability.
<sup>b</sup>: In the case of the RIF+CLA+AMX/CLV combination, the shift was estimated relative to the *apparent* potency of RIF + CLA, i.e., $EC_{50_{combo}} = EC_{50_{RIF}} * \Theta_{EC50_{CLA}} * \Theta_{EC50_{CLA+AMX/CLV}}$
<sup>c</sup>: Residual unexplained variability was modelled as additive in the logarithm scale.

For clarity, **Figure S*4*** provides an overview of the final drug-disease model describing the effect of drug-combinations in vitro. Mathematical equations for the model are included in the Appendix.

### 3.2. Clinical trial simulations

Parameter estimates of the final drug-disease model were used to simulate antibacterial activity in conjunction with clinical PK models, taking into account interindividual variability in pharmacokinetic properties of the drugs, and assuming comparable bacterial growth dynamics and PKPD relationships *in vitro* and *in vivo*, in a clinical setting with paediatric and adult patients affected by BU. Simulated median skin Css values in patients for the RIF+CLA+AMX/CLV arm were predicted to be 0.14 µg/mL, 0.22 µg/mL and 0.71 µg/mL for RIF, CLA and AMX, respectively (***Figure 5***), with a variability (standard deviations normalised to the median Css) amounting to 53% for RIF, 35% for CLA, and 49% for AMX. These results indicated substantial variability in exposure to the three antimicrobials, which might significantly influence the overall pharmacological efficacy of the combination treatments in the simulated populations. Simulated steady-state systemic PK profiles for all treatment arms (n = 70 subjects per arm) are presented in **Figure S*5***.

**Figure 5:**
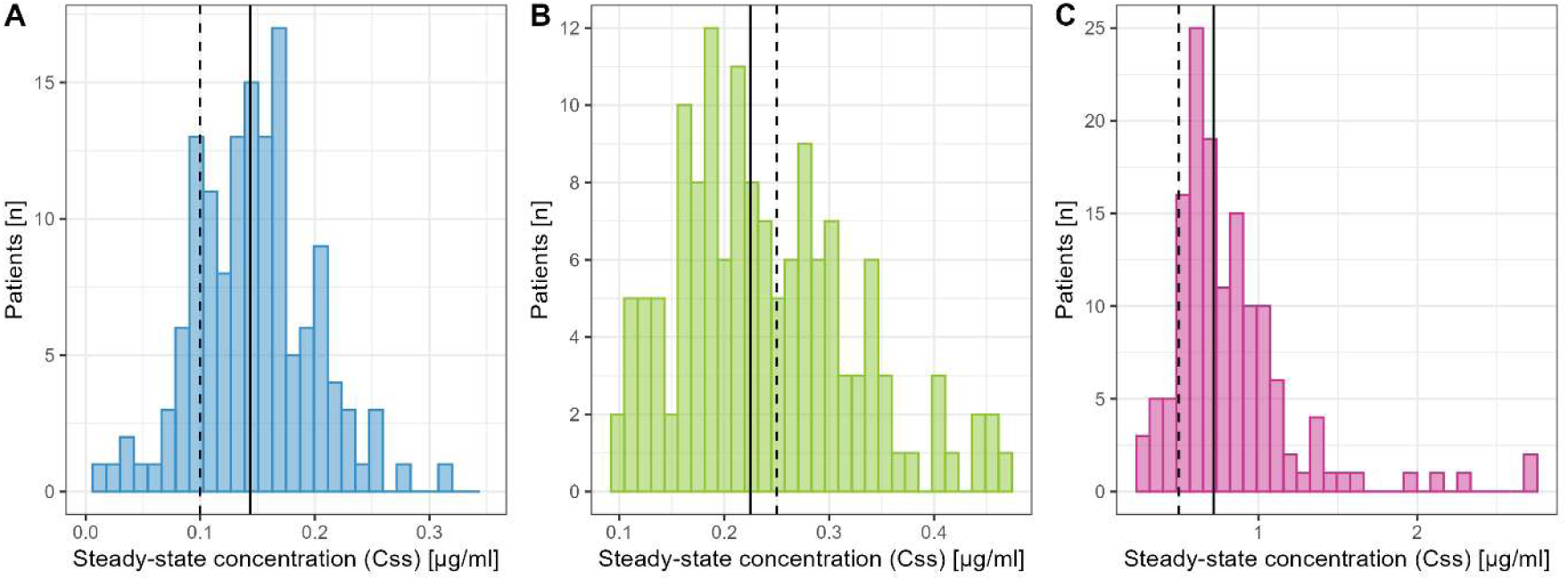
Predicted steady-state concentrations (Css) in the skin based on simulation of the individual pharmacokinetic profiles of virtual cohort of patients (n=70). (A) Rifampicin (q.d., 10mg/kg), (B) Clarithromycin (b.i.d., 7.5 mg/kg), (C) Amoxicillin (b.i.d., 1000 mg/kg). Solid lines represent the median value of the distribution, while dashed lines represent the *in vitro* MIC of the drug, which also correspond to the highest concentration tested in combination experiment.

Figure 6 and **Table S*3*** show the predicted clinical bacterial eradication rates for the investigated treatments for each clinical isolate, stratified by baseline bacterial load. The bacterial burden at the start of treatment notoriously emerged as the primary factor influencing the time to bacterial eradication, irrespectively of treatment choice. For initial bacterial loads up to 1,000 CFU/ml, most patients achieved eradication at 28 days in all simulated treatment arms. In fact, all patients with an initial bacterial load of 10 CFU/mL achieved eradication in 14 days, and in 21 days for an initial bacterial load of 100 CFU/mL in almost 100% patients with the exception of the clinical isolate from China. However, simulated patients with an initial bacterial load of 10,000 CFU/mL typically required more than 28 days to achieve sterilisation, with some needing over 56 days, depending on bacterial susceptibility, drug combination, regimen and overall drug exposure. In general, AMX/CLV-containing regimens outperformed the standard of care in terms of bacterial eradication rates, with significant differences at higher initial bacterial load (1,000-10,000 CFU/mL). Notably, simulations involving strain ITM070290 showed the need of markedly increased times to eradication; in the case of high initial bacterial loads (10,000 CFU/ml), a significant number of patients (more than 20%) would require more than 8 weeks of antibiotic treatment for eradication, partly explaining recurrence of the infection in some patients following treatment according to current guidelines.

**Figure 6:**
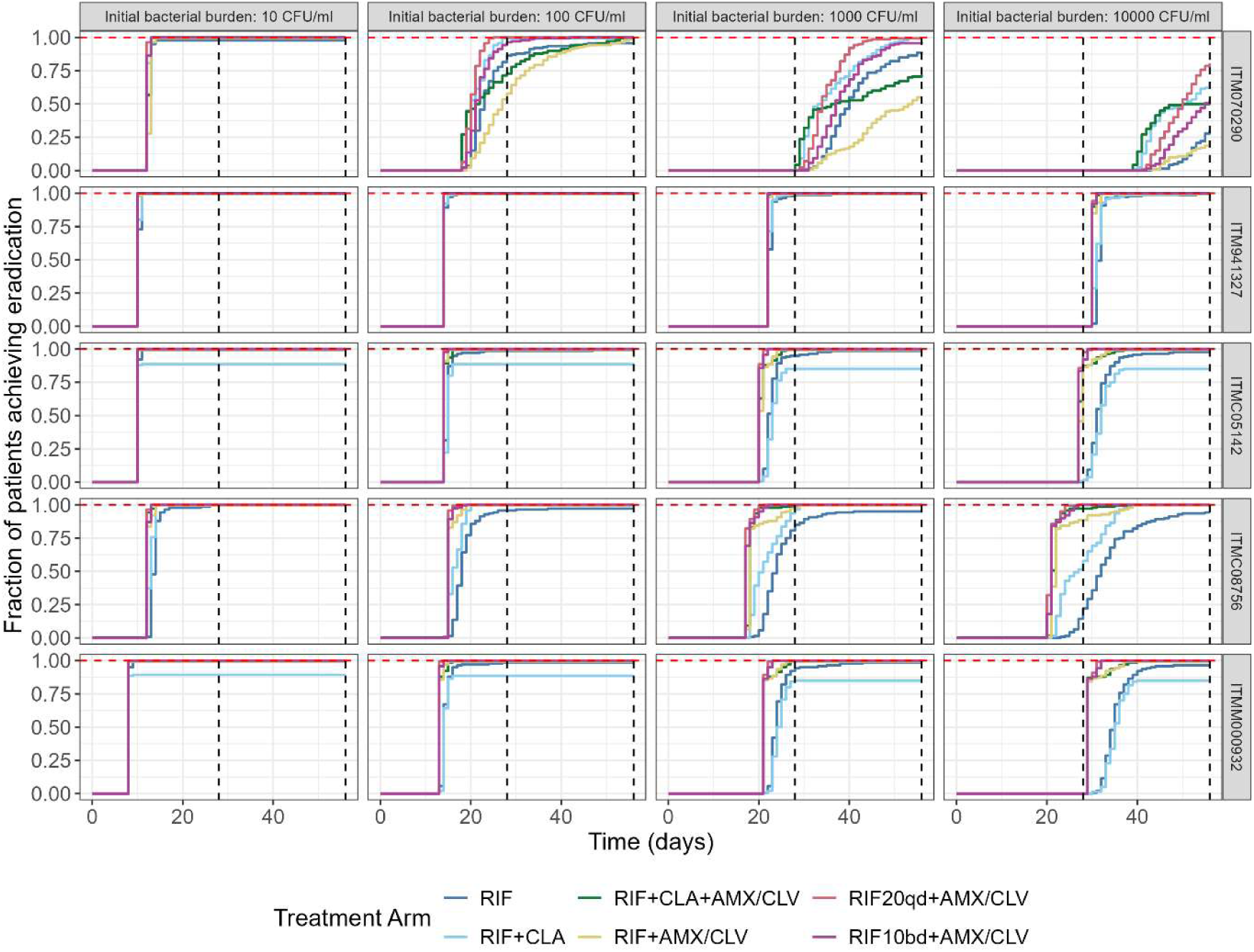
Fraction of simulated patients achieving bacterial eradication over time for different investigational treatments, initial bacterial burden and bacterial susceptibility. Simulation scenarios take into account the variability in pharmacokinetic properties within the simulated population. Columns represent scenarios with different bacterial load at the start of the treatment, while rows depict results for five different clinical isolates. Black dashed lines represent 4 and 8 weeks of treatment, while the red dashed line represents complete eradication in the simulated cohort of patients.

## 4. Discussion and conclusions

The current BU standard of care treatment, recommended by the WHO in 2017, achieves a cure rate of approximately 96%^7^. However, this regimen is based on a 8-week course of RIF and CLA compromises patient adherence, particularly in rural or economically disadvantaged areas, where regular access to health care facilities may be difficult. Patients may also face difficulties adhering to the full course of treatment due to various socioeconomic factors, including transportation, school drop-out, loss of income during treatment or lack of family support. Treatment also includes extensive wound care, sometimes surgical intervention, and physiotherapy rehabilitation of movement limitations and disability, with lesion healing potentially lasting for several months. Consequently, the strategic direction of the WHO targets the development of novel regimens with the potential to reduce the duration of treatment^2^.

On the other hand, there is an ongoing debate whether current antimicrobial BU treatments (RIF+CLA) would allow for eradication of *M. ulcerans* from the lesions in only 4 weeks. Previous clinical observations suggest that a 4-week regimen of RIF plus streptomycin combination— against which the current standard of care RIF + CLA has been shown to be non-inferior^7^— results in complete bacterial clearance in early BU lesions (plaques and nodules). Similarly, Sarpong-Duah and colleagues^30^ found that the standard of care could achieve sterilisation in lesion that were associated with a low baseline bacterial load. These results were contrasted by *in vivo* investigations performed by Almeida et al.^31,32^, who evaluated several drug combinations in a murine footpad infection model. With a disease incubation period of 35 days, they reported that RIF (10 mg/kg q.d.) and RIF+CLA (10 mg/kg q.d. and 100 mg/kg q.d., respectively) showed similar bactericidal activity in mice, reaching sterilisation at 6 or 8 weeks. Even with a shorter incubation period of 11 days, those regimens could not reach sterilisation within 4 weeks, although it must be noted different *M. ulcerans* strains were used in the two studies and that the pathology in the mouse footpad infection model might not fully reflect the human disease.

Historically, the selection of novel regimens and companion drugs to be used in investigational antimicrobial combinations, as well as the overall treatment duration, have been decided empirically^13^. As such an approach disregards the underlying PKPD relationships, empirical choices have often resulted in treatment failure^33,34^ . This issue also applies to BU, for which some treatments currently in use have been selected empirically^5,35^, despite the possibility of a systematic rationale for the translation of preclinical findings to inform and/or prioritise choices regarding companion drugs and dosing regimen in the clinic. In fact, we have previously attempted show the value of a translational framework to select and justify the dose rationale in the clinic using a model-based approach to link *in vitro* MIC values with the PTA in humans^11^. Here we expand on the same principles to include bacterial growth dynamics using data from *in vitro* time kill assays with clinical isolates of *M. ulcerans*. Assuming that clinically relevant pharmacodynamic drug interactions can be parameterised as an increase potency, PKPD models have been developed that provide insight into bacterial clearance and time to eradication taking into account varying susceptibility of different clinical isolates. Together with clinical trial simulations, this approach offers an opportunity to also characterise the implications of interindividual differences in pharmacokinetics and drug distribution to the site of infection, including the effect of doses, dosing regimens and treatment duration.

Our results rely on a semi-mechanistic translational framework that predicts antibacterial activity of different regimens and drug combinations for BU, borrowing concepts from established approaches in TB research^13,22^. Of note is the finding that the baseline bacterial load largely determines the time to bacterial eradication, as shown in prior studies^30,36^. Our simulation scenarios also show that bacterial eradication is predicted to be achieved in 28 days in patients with low bacterial loads at the start of treatment (10 - 100 CFU/mL), regardless of the selected regimen. By contrast, antibacterial regimen choice and interindividual variability in drug exposure in lesions become pivotal factors determining treatment success in patients with higher bacterial load. In fact, our findings show that regimens including AMX generally outperform the standard of care in patients with initial bacterial loads of 1,000 CFU/ml; these regimens were capable of achieving consistent bacterial eradication within 4 weeks. However, the ITM070290 isolate represented an important exception in our analysis, with markedly longer predicted time to eradication, due to a lower susceptibility to RIF and less pronounced effect of the combination with AMX/CLV.

It is worth mentioning that our results align with previous reports highlighting the strong *in vitro* synergy of AMX/CLV with RIF^8,9^, and its enhanced PTA in the clinic^11^. Of note, simulated regimens featuring the combination of RIF at high doses (either 20 mg/kg q.d. or 10 mg/kg b.i.d.) and AMX/CLV performed close, or in some cases slightly superior, to the triple drug combination with RIF at standard doses. This raises the possibility of substituting CLA with AMX/CLV entirely, thus maintaining an equally, or more effective simplified regimen. Clinical experience in TB research has shown that increasing RIF doses up to daily 35 mg/kg is safe^17^; thus, adopting a regimen of 20 mg/kg once daily or 10 mg/kg twice daily is not expected to add significant burden on patients in terms adherence and safety. A twice-daily administration of RIF would also align conveniently with the twice daily dosing schedule of AMX/CLV. Furthermore, AMX is not subject to markedly reduced clinical exposure due to the RIF-mediated induction of CYP450 metabolism, a known limitation of CLA^37,38^ which was explicitly considered in our simulations. Even though our model does not include the limited inhibitory effect of CLA on CYP450, which counteracts some of the auto-induction observed in RIF metabolism, overall our results suggest that CLA could be replaced by AMX/CLV, without further clinical implications for the antibacterial activity.

While our work provides insight into how *in vitro* preclinical data can be integrated to estimate the likelihood of pharmacological success in clinical settings, there are limitations to consider. Firstly, TKA with RIF, CLA, and AMX/CLV combinations were tested only at two matching concentration levels (0.25xMIC and 1×MIC). Consequently, assumptions had to be made to characterize PD interactions between these drugs under those specific conditions. A pragmatic approach was used to describe the potential effect with varying concentrations in vivo. We have assumed that similar interactions would occur in the clinical setting when simulated individual drug concentrations approximated those tested *in vitro*. Secondly, the linear relationship between RLU and CFU used in our simulations was derived from a relatively sparse dataset and did not cover the lower range of CFU/mL and RLU values (i.e., below 10⁴) which we simulated within our analysis. Nevertheless, linearity between RLU and CFU/mL biomarkers is well established in the literature and previous studies indicate that the limit of detection for RLU using the Bactiter Glo assay is approximately 10³–10⁴ CFU/mL, which can make generating robust data in the lower range technically challenging. Importantly, evidence from a study on *Mycobacterium tuberculosis* suggests that the linear relationship may extend down to 10² CFU/mL^24^, supporting the plausibility of our assumption of linearity even at lower bacterial burden. Lastly, a significant factor missing from our analysis was the immune system’s contribution to bacterial clearance. It is well-established that a hallmark of BU pathophysiology is the production of mycolactone by *M. ulcerans,* a toxin which distributes extensively both at the site of infection and systemically, causing a downregulation of multiple mediators of inflammation^39,40^. As antibiotic treatment begins and mycolactone levels lower due to bacterial death, immune system functionality is progressively restored^,48^. Nonetheless, the extent to which the restored immune-mediated clearance contributes to cure in the clinic, and how variable this effect is, remains unclear since recurrence is typically observed after a variable period of time^43,44^. Considering that the antimicrobials tested in the work do not exhibit immunomodulatory properties, our simulations might underestimate the time to eradication across tested regimens, especially in high baseline bacterial load scenarios, cases in which the immune system might exert its containing effect. This could be especially relevant for AMX/CLV-containing combinations and the high dose RIF regimen, positioning them as even stronger candidates to shorten the treatment to 28 days.

Finally, clinical factors that influence the rate of wound healing, such as the extent of the lesion and presence of co-infection were not included in our simulations^36^. Clinical cure in BU is typically defined by wound healing, not solely by bacterial eradication. Proper wound care is then critical to promote timely healing, prevent secondary infections, and minimize the risk of scarring or disability^45^. On the other hand, addition of AMX/CLV at the proposed doses and dosing regimen could act as prophylactic to secondary infections due to *Staphylococcus aureus*, improving wound care.

In summary, we have established a translational framework to evaluate the probability of pharmacological success of investigational BU treatments leveraging *in vitro* longitudinal time-kill data. As the triple-drug regimen (RIF+CLA+AMX/CLV) was predicted to achieve complete bacterial eradication within 4 weeks in most scenarios, our simulations support the BLMs4BU clinical trial^10^ (*NCT05169554*, *PACTR202209521256638)*, in which the addition of AMX/CLV to the standard of care is being evaluated to shorten the treatment for BU from 8 to 4 weeks. Notably, simulation scenarios with RIF+AMX/CLV combinations were predicted to achieve comparable success rates. This regimen could open an avenue for further simplification of BU treatment, while potentially minimizing secondary side effects associated to the use of CLA.

## Data Availability

All data produced in the present study are available upon reasonable request to the authors

