## Supplementary Material for "Prediction of Buruli ulcer treatment shortening with novel beta-lactam-containing antimicrobial combinations"

### Appendix A: model equations

The final *in vitro* drug–disease model describing the effect of the antimicrobial combinations incorporates delayed logistic bacterial growth and a first-order bacterial killing process characterized by an Emax model. The system is described by the following differential equation:

$$\frac{dB}{dt} = \left( GR \cdot \left( \frac{B(t-\tau)}{B_{max}} \right) - \; H(t - T_{lag}) \cdot \frac{Emax_{RIF} \cdot C_{rif}(t)}{EC_{50} + C_{rif}(t)} \right) * B(t)$$

Here, $B(t)$ represents the mycobacterial burden at time t, expressed as RLU values. $GR$ denotes the maximal mycobacterial growth rate, $B_{max}$ the carrying capacity of the system, $\tau$ the delay term describing the dependency of growth on past bacterial burden. $H\left(t - T_{lag}\right)$ is a step function that defines the onset of the drug effect, such that the Emax-driven killing is initiated only after a lag time $T_{lag}$. $C_{rif}(t)$ is the concentration of rifampicin at time t, and $Emax_{RIF}$ represents the maximum bacterial killing rate of RIF. When RIF is administered as monotherapy, $EC_{50}$ reflects its intrinsic potency. In the presence of a companion drug, $EC_{50}$represents the apparent potency of the combination and is defined as:

$$EC_{50} = \begin{cases} EC_{50_{RIF}} & if\ RIF\ alone \\ EC_{50_{RIF}} * \Theta_{EC50_{combo}} & if\ combo \end{cases}$$

Where $\Theta_{EC50_{combo}}$ is a multiplicative shift parameter quantifying the effect of the tested drug combination on rifampicin potency.

### Appendix B: linear relationship between CFU and RLU

For a subset of the relative light unit (RLU) time-kill assay (TKA) samples used in this study, the corresponding quantification of colony-forming units (CFUs) was available. Both RLU and CFU values were log-transformed, and a linear regression model was fitted in R to establish a relationship between the two quantities. The final regression parameters, as well as the final regression line, are depicted in **Figure S1**.

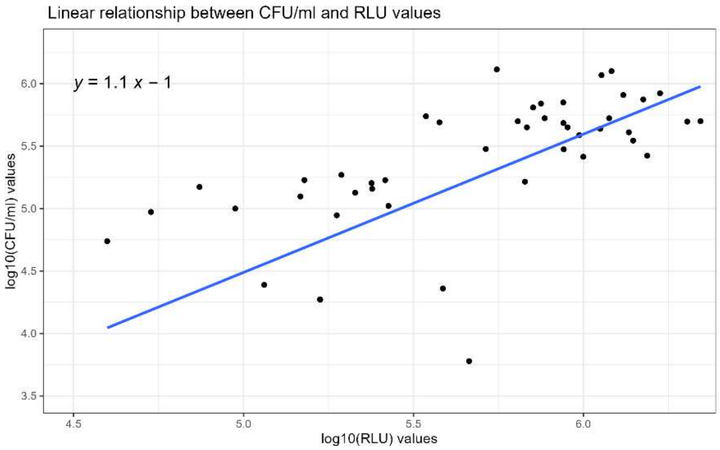


**Figure S1: Linear regression describing the correlation between log-transformed CFU/ml and RLU values.** Individual dots represent experimental samples for which both markers of viability were available, and the blue line represent the linear trend. The equation on the top-left side of the graph represents the final regression line.

### Supplementary Figures

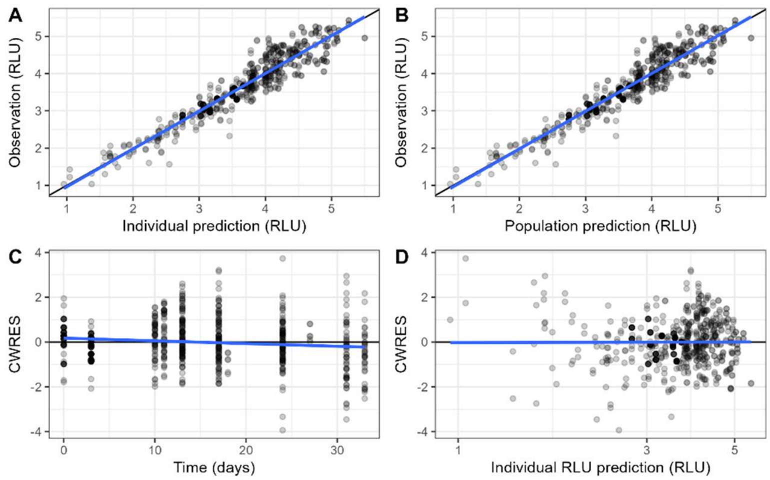


**Figure S2: Goodness-of-fit plots for the drug-disease model describing the natural growth of *Mycobacterium ulcerans* and the monotherapy concentration-effect relationship of RIF *in vitro*.** A) Observations *vs* individual predictions plot; B) Observations *vs* population predictions plot; C) CWRES *vs* time plot; D) CWRES *vs* individual predictions plot. In each subgraph, the solid blue line represents the linear regression line built from the scatterplot. Abbreviations: CWRES = conditional weighted residuals; RLU = relative light units.

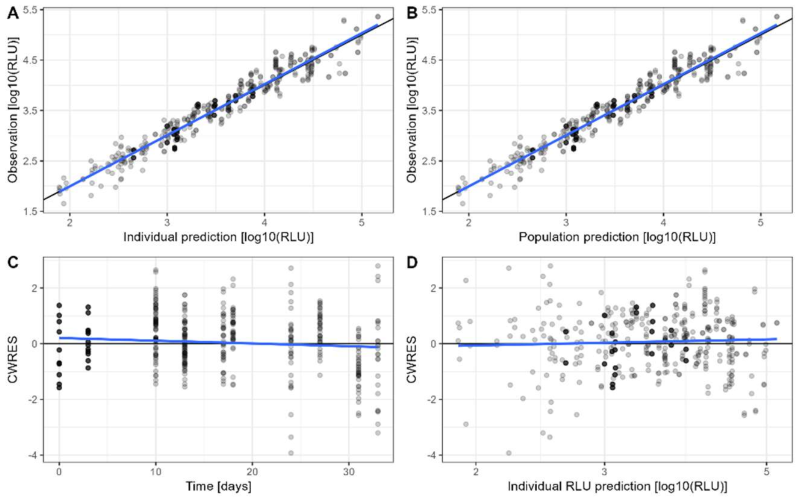


**Figure S3: Goodness-of-fit plots for the drug-disease model describing the natural growth of *Mycobacterium ulcerans* and the effect of drug combinations *in vitro*.** A) Observations *vs* individual predictions plot; B) Observations *vs* population predictions plot; C) CWRES *vs* time plot; D) CWRES *vs* individual predictions plot. In each panel, the solid blue line represents the linear regression line built from the scatterplot. Abbreviations: CWRES = conditional weighted residuals; RLU = relative light units.

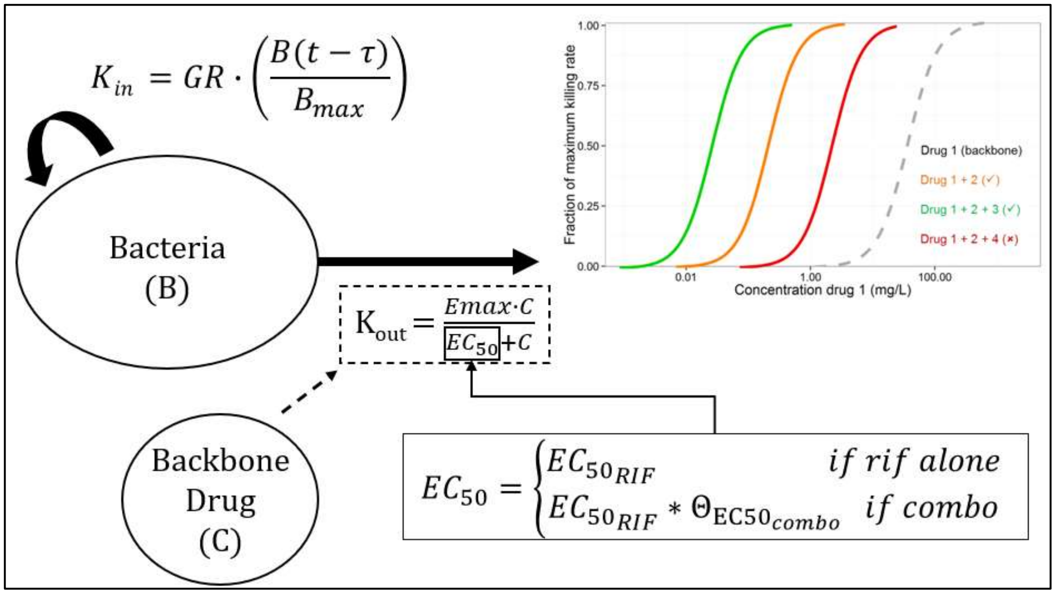


**Figure S4: Schematised representation of the final model describing the effect of drug combination in vitro.** A delay logistic model describes the natural growth of bacteria ($K_{in}$), while bacterial killing ($K_{out}$) follows a first-order process described by an Emax model coupled with delayed onset of effect ($T_{lag}$). In absence of companion drugs, the overall potency corresponds to that of rifampicin alone. When companion drugs are present, the overall *apparent* potency is modified by a multiplicative shift specific to each companion drug and its concentration. The graph exemplifying the shift of the concentration-effect curve of the backbone drug was reproduced with permission from Muliaditan et al. Abbreviations: GR = maximal replication rate for M. ulcerans, $B_{max}$ = carrying capacity of the system; τ = time dependence of bacterial replication; C = drug concentration; Emax = Maximal elimination rate of the backbone drug (RIF); $EC_{50}$ = concentration of the backbone drug at which half-maximal elimination rate is achieved; $\Theta EC_{50\_combo}$ = shift to the potency of the backbone drug due to companion drugs

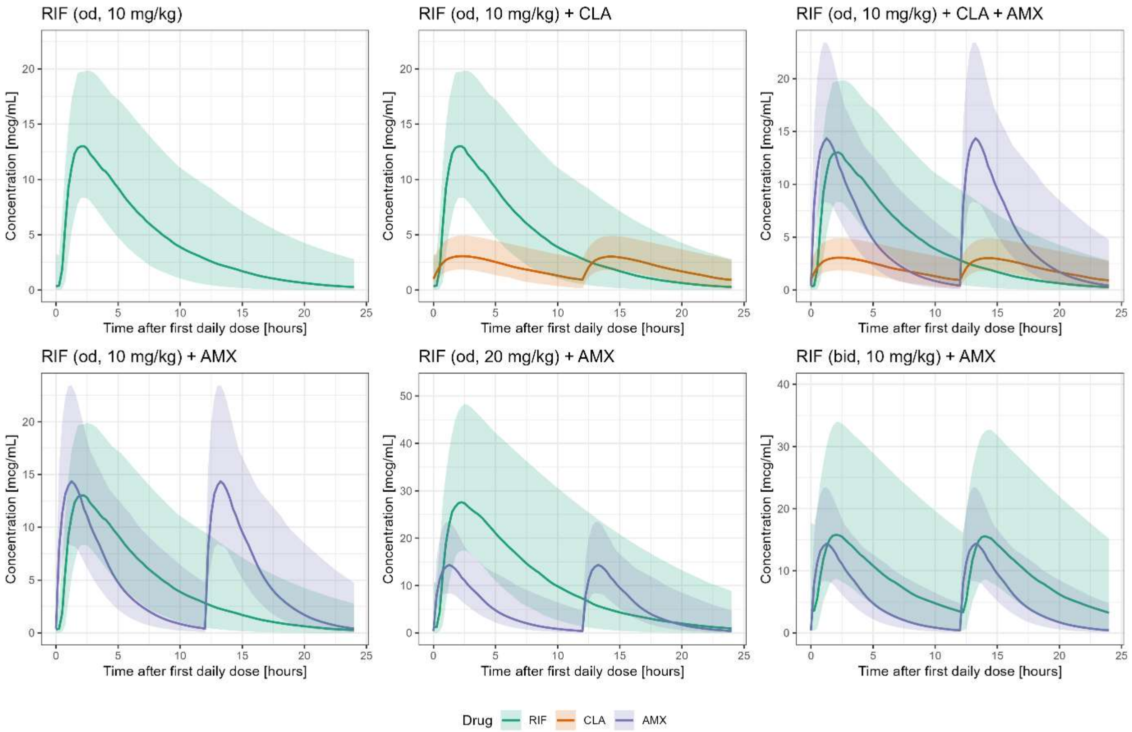


**Figure S5: Simulated systemic concentration-time profiles for each drug (RIF, CLA and AMX) at steady state for the investigated treatment arms.** Each arm includes n = 70 virtual patients. Solid lines represent the median pharmacokinetic (PK) profiles, and the surrounding shaded areas indicate the 90% confidence interval around the median. Abbreviations: RIF = rifampicin, CLA = clarithromycin, AMX = amoxicillin

### Supplementary Tables

**Table S1: Population demographics in West African children, taken from a cross sectional study of children aged 6 to <18 years in Calabar, South Nigeria.**

| MALES | | | | FEMALES | | | |
|---|---|---|---|---|---|---|---|
| **AGE (YEAR)** | Mean Weight (kg) | Mean Height (cm) | BMI ($kg/m^2$) | Age (year) | Mean Weight (kg) | Mean Height (cm) | BMI ($kg/m^2$) |
| **6** | 21.9 | 117.6 | 15.7 | 6 | 20.9 | 116.0 | 15.5 |
| **7** | 26.8 | 126.8 | 16.6 | 7 | 26.1 | 128.2 | 15.8 |
| **8** | 28.9 | 131.9 | 16.6 | 8 | 27.6 | 129.8 | 16.2 |
| **9** | 31.0 | 136.5 | 16.6 | 9 | 34.9 | 138.1 | 18.0 |
| **10** | 34.2 | 140.1 | 17.5 | 10 | 34.8 | 143.1 | 16.9 |
| **11** | 36.0 | 144.5 | 17.1 | 11 | 39.4 | 146.8 | 18.0 |
| **12** | 38.9 | 148.1 | 17.6 | 12 | 43.2 | 152.5 | 18.6 |
| **13** | 42.6 | 153.3 | 17.9 | 13 | 45.7 | 154.2 | 19.0 |
| **14** | 45.5 | 157.8 | 18.2 | 14 | 50.9 | 156.8 | 20.6 |
| **15** | 50.7 | 160.8 | 19.7 | 15 | 52.0 | 159.2 | 20.2 |
| **16** | 57.1 | 167.6 | 20.3 | 16 | 51.6 | 156.2 | 21.2 |
| **17** | 57.2 | 168.3 | 20.1 | 17 | 51.6 | 155.1 | 21.4 |
| **18** | 58.2 | 165.6 | 21.2 | 18 | 61.5 | 161.5 | 23.5 |

**Table S2: Pharmacokinetic parameters relating total concentrations in plasma to drug concentrations in the skin.** The plasma-to-skin ratio was incorporated as a scaling factor into the final PKPD model to describe skin exposure following oral doses of rifampicin, clarithromycin and amoxicillin. Tissue equilibration was assumed to be achieved concurrently with steady state conditions in plasma. Scarring and fibrotic tissue were considered to have minor effect on drug permeability and tissue perfusion.

| Parameter | Value (%) | Reference | Notes |
|---|---|---|---|
| **Rifampicin** | | | |
| unbound conc. | 9 | Litjens *et al.*[50] | |
| unbound conc. in plasma to skin ratio | 68 | Kenny *et al.*[51] | |
| **Clarithromycin** | | | |
| unbound conc. | 30 | Rodvold[52] | |
| unbound conc. in plasma to skin ratio | 39 | Traunmüller *et al.*[53] | |
| **Amoxicillin** | | | |
| unbound conc. | 80 | Huttner *et al.*[54] | |
| unbound conc. in plasma to skin ratio | 17 | Shukla[55] | No data was available in humans, the reported value comes from a preclinical study in rabbits. |

**Table S3: Predicted proportion of patients achieving bacterial eradication after 4, 6 and 8 weeks of treatment, stratified by antimicrobial regimen, initial bacterial load in the lesion and *Mycobacterium ulcerans* strain.** Results are shown for the two highest baseline bacterial load scenarios.

| Treatment arm | Probability of bacterial eradication (%) | | | | | |
|---|---|---|---|---|---|---|
| | Baseline bacterial load: 1000 CFU/ml | | | Baseline bacterial load 10000 CFU/ml | | |
| | 4 weeks | 6 weeks | 8 weeks | 4 weeks | 6 weeks | 8 weeks |
| ITM070290 | | | | | | |
| RIF | 0 | 53.6 | 85 | 0 | 0 | 30 |
| RIF+CLA | 0 | 81.4 | 100 | 0 | 22.1 | 66.4 |
| RIF+CLA+AMX/CLV | 4.3 | 52.9 | 72.1 | 0 | 31.4 | 50.7 |
| RIF+AMX/CLV | 0 | 22.9 | 55 | 0 | 0.7 | 22.1 |
| RIF20qd+AMX/CLV | 0.7 | 95 | 99.3 | 0 | 4.3 | 80.7 |
| RIF10bid+AMX/CLV | 0 | 80 | 96.4 | 0 | 1.4 | 52.9 |
| ITM941327 | | | | | | |
| RIF | 98.6 | 100 | 100 | 0 | 98.6 | 100 |
| RIF+CLA | 100 | 100 | 100 | 0 | 98.6 | 100 |
| RIF+CLA+AMX/CLV | 100 | 100 | 100 | 0 | 98.6 | 100 |
| RIF+AMX/CLV | 100 | 100 | 100 | 0 | 100 | 100 |
| RIF20qd+AMX/CLV | 100 | 100 | 100 | 0 | 100 | 100 |
| RIF10bid+AMX/CLV | 100 | 100 | 100 | 0 | 100 | 100 |
| ITMC05142 | | | | | | |
| RIF | 95 | 98.6 | 98.6 | 1.4 | 96.4 | 97.9 |
| RIF+CLA | 85 | 85 | 85 | 1.4 | 85 | 85 |
| RIF+CLA+AMX/CLV | 100 | 100 | 100 | 87.1 | 100 | 100 |
| RIF+AMX/CLV | 100 | 100 | 100 | 86.4 | 100 | 100 |
| RIF20qd+AMX/CLV | 100 | 100 | 100 | 97.9 | 100 | 100 |
| RIF10bid+AMX/CLV | 100 | 100 | 100 | 92.1 | 100 | 100 |
| ITMC08756 | | | | | | |
| RIF | 79.3 | 94.3 | 96.4 | 22.1 | 87.1 | 94.3 |
| RIF+CLA | 96.4 | 100 | 100 | 57.9 | 100 | 100 |
| RIF+CLA+AMX/CLV | 98.6 | 100 | 100 | 97.1 | 100 | 100 |
| RIF+AMX/CLV | 97.9 | 100 | 100 | 88.6 | 100 | 100 |
| RIF20qd+AMX/CLV | 100 | 100 | 100 | 100 | 100 | 100 |
| RIF10bid+AMX/CLV | 100 | 100 | 100 | 100 | 100 | 100 |
| ITMM000932 | | | | | | |
| RIF | 94.3 | 97.9 | 98.6 | 0 | 94.3 | 96.4 |
| RIF+CLA | 85 | 85 | 85 | 0 | 85 | 85 |
| RIF+CLA+AMX/CLV | 100 | 100 | 100 | 0 | 100 | 100 |
| RIF+AMX/CLV | 100 | 100 | 100 | 0 | 100 | 100 |
| RIF20qd+AMX/CLV | 100 | 100 | 100 | 0 | 100 | 100 |
| RIF10bid+AMX/CLV | 100 | 100 | 100 | 0 | 100 | 100 |
